# Progress of the Delta variant and erosion of vaccine effectiveness, a warning from Utah

**DOI:** 10.1101/2021.08.09.21261554

**Authors:** Lindsay T. Keegan, Shaun Truelove, Justin Lessler

## Abstract

Since the emergence of SARS-CoV-2, vaccines have been heralded as the best way to curtail the pandemic. Clinical trials have shown SARS-CoV-2 vaccines to be highly efficacious against both disease and infection. However, those currently in use were primarily tested against early lineages. Data on vaccine effectiveness (VE) against variants of concern (VOC), including the Delta variant (B.1.617.2), remain limited. To examine the effectiveness of vaccination in Utah we compared the proportion of cases reporting vaccination to that expected at different VEs, then estimated the combined daily vaccine effectiveness using a field evaluation approach. Delta has rapidly outcompeted all other variants and, as of June 20th, represents 70% of all SARS-CoV-2 viruses sequenced in Utah. If we attribute the entire change in VE to the Delta variant, the estimated vaccine effectiveness against Delta would be 82% (95% CI: 78%, 85%). We show a modest reduction in vaccine effectiveness against COVID-19 in Utah corresponding to the expansion of the Delta lineage in the state. This reduction in the effectiveness of available vaccines correlated with the arrival of novel VOCs, rather than waning immunity, is highly concerning.

## Introduction

Since the emergence of SARS-CoV-2, vaccines have been heralded as the best way to curtail the pandemic. Clinical trials have shown SARS-CoV-2 vaccines to be highly efficacious against both disease and infection [1]. However, those currently in use were primarily tested against early lineages. Data on vaccine effectiveness (VE) against variants of concern (VOC), including the Delta variant (B.1.617.2), remain limited.

The spread of VOCs with significantly higher transmissibility (e.g., Delta), have raised concerns about ability for vaccines to sustainably control SARS-CoV-2. These concerns are exacerbated by increasing numbers of cases reported in fully vaccinated individuals as the Delta variant spreads globally; potentially signaling a decreased VE for VOCs [2]. A preliminary study in the UK on the VE of the Pfizer/BioNTech and AstraZeneca vaccines found 6% reduction in efficacy for Pfizer and 12% for AstraZeneca [3]. However, because of Delta’s recent emergence and low case numbers, this study only included 27 fully vaccinated cases altogether.

## Methods

The Utah Department of Health (UDOH) began monitoring VOCs in October 2020 and first identified two cases infected with Delta in mid-April 2021[4]. UDOH currently sequences 10% of reported COVID-19 cases and collects vaccination coverage for all cases. Data used for this analysis included unlinked daily numbers for vaccination status and proportion of variant among cases. Its emergence, Delta has rapidly outcompeted all other variants and, as of June 20^th^, represents 70% of all lineages sequenced in Utah (Figure 1C). Simultaneously, the percent of breakthrough cases in Utah was increasing at a concerning rate (Figure 1A). However, increasing vaccination coverage is expected to result in increased frequency of breakthrough infections, even if VE remains constant (Figure 1A). To determine the effectiveness of the vaccines deployed in Utah and investigate the rate of breakthrough infections in the face of a rapidly changing variant landscape, we estimated the combined daily vaccine effectiveness (*VE*_*t*_) using a field evaluation approach developed by Orenstein et al. [5]. To estimate the VE against Delta alone, we partition *VE*_*t*_ into the VE of the Delta variant (*VE*_*δ*_) and the VE of all other variants (*VE*_*1-δ*_), such that:

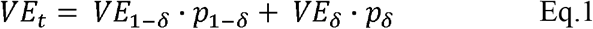

**Figure 1.**
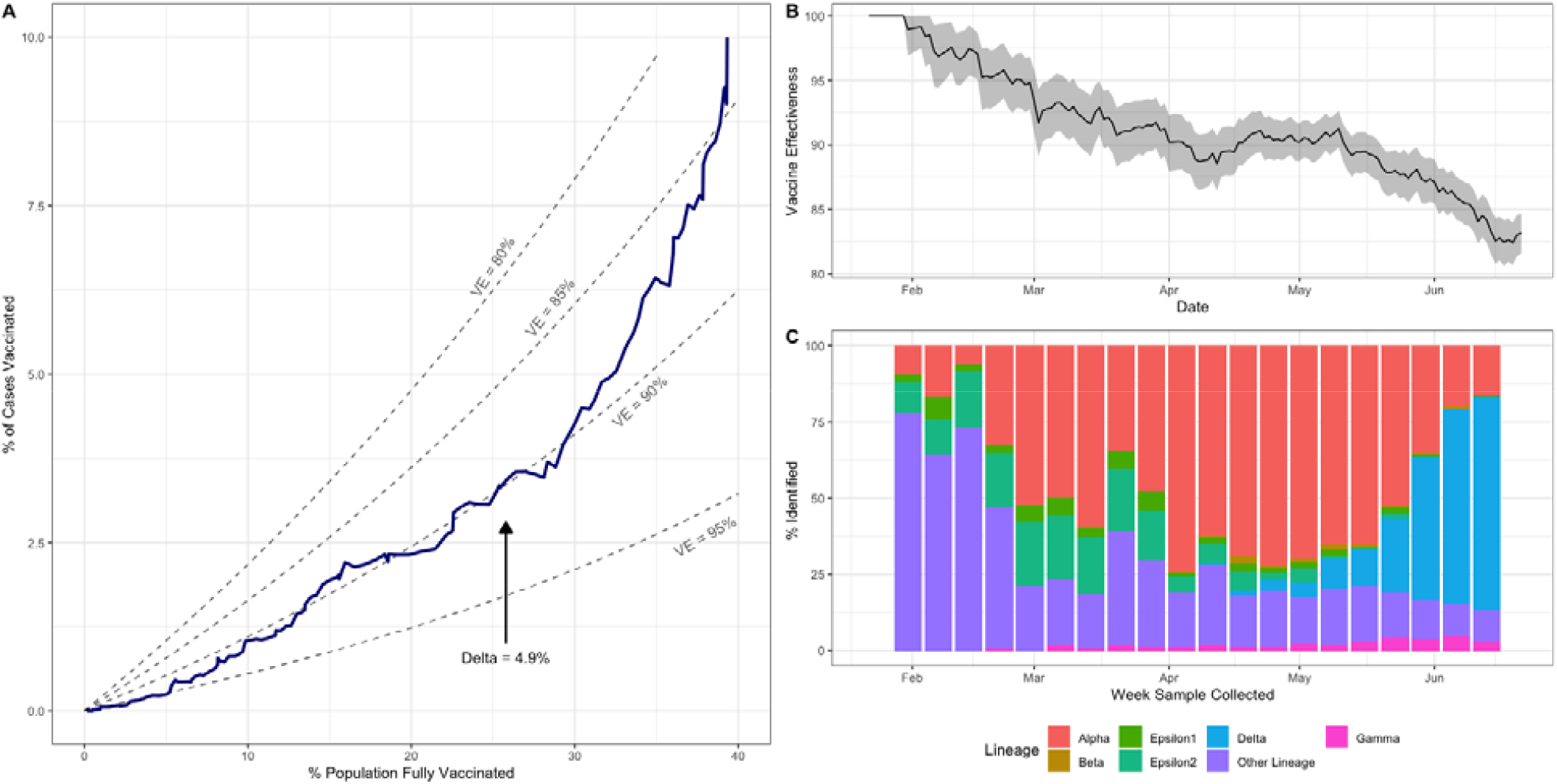
Daily percent of breakthrough cases and population that is fully vaccinated, daily vaccine effectiveness against SARS-CoV-2, and sequencing results by week, in Utah. (A) The 14-day moving average of the percent of breakthrough cases against the percent of the population vaccinated (blue) from January 16, 2021 to June 28, 2021 with the theoretical curves for expected percent of breakthrough cases with vaccine effectiveness ranging from 80% to 95% (grey dashed lines). (B) 14-day moving average of vaccine effectiveness against SARS-CoV-2 from January 16, 2021 to June 28, 2021 and the 95% confidence interval (shaded grey region). (C) The weekly percent of all sequenced samples by lineage from the week starting on January 17, 2021 to the week starting on June 13, 2021.

## Results

As of June 28, 2021, 54% of the eligible population (1,407,233 of 3,205,958 Utahans) were fully vaccinated (defined as 14+ days post final vaccine dose); 52.9% with Pfizer, 38.1% with Moderna, and 9.05% with Janssen [6]. The proportion of breakthrough cases started increasing faster than expected based on population vaccination rates in mid-May (Figure 1A), reaching 10.5% by the end of June, versus 6.4% as expected with a 90% effective vaccine. We estimated the effective VE from all vaccination in Utah from January 16 – June 28, 2021 (Figure 1B), and find that VE declines from 90% in mid-May to 83% by the end of June (Figure 1B). This decline occurs at the same time as a rapid increase in the proportion of cases infected with the Delta variant (Figure 1C). The first identified case in Utah infected with Delta occurred in mid-April 2021[4]. Since then, Delta has rapidly outcompeted all other variants and, as of June 20^th^, represents 70% of all SARS-CoV-2 viruses sequenced in Utah (Figure 1C). If we attribute the entire change in VE to the Delta variant (i.e., *VE*_*t*_ *= VE*_*1-δ*_ *· p*_*1-δ*_ *+ VE*_*δ*_ *· p*_*δ*_), the estimated vaccine effectiveness against Delta would be 82% (95% CI: 78%, 85%).

## Discussion

We show a modest reduction in vaccine effectiveness against COVID-19 in Utah corresponding to the expansion of the Delta lineage in the state. This reduction in the effectiveness of available vaccines correlated with the arrival of novel VOCs, rather than waning immunity, is highly concerning. This should serve as a caution to states throughout the US that the Delta variant has the potential to bring with it renewed outbreaks, even in highly vaccinated populations. If there is a consistent trend of increasing immune escape as new variants arise, it could eventually undermine the effectiveness of current vaccines and necessitate mass re-vaccination.

## Limitations

We used publicly available data, collected for outbreak surveillance, which has limitations. The breakthrough case definition allows for self-reporting of vaccination status which may result in fully vaccinated individuals reporting at higher rates (10% overreporting of vaccination would bring the June VE to 85.0%). Also, each day only 10-15% of cases were sequenced with priority to breakthrough cases and outbreaks, potentially causing an oversampling of Delta variants. Additionally, we do not account for known demographic differences (e.g., urbanicity) in unvaccinated and breakthrough infections.

## Data Availability

All data are publicly available from the Utah Department of Health.

https://coronavirus.utah.gov/case-counts/

## Acknowledgements

We would like to acknowledge the Utah Department of Health for their tireless efforts to collect and share data on SARS-CoV-2 infection, vaccination, and sequencing. LTK was supported by the Centers for Disease Control and Prevention (grant nos. 5U01CK000585-02 and 5U01CK000555-02). ST report funding from a NSF COVID-19 RAPID award and ST and JL from the U.S. Department of Health and Human Services (DHHS), Office of the Assistant Secretary for Preparedness and Response to the Johns Hopkins Applied Physics Laboratory.

<u>COI:</u> LTK has worked with Pfizer modeling the impact of vaccines in long-term care facilities. JL has worked as an expert witness on cases where the length and severity of the COVID-19 pandemic are of issue. ST has consulted on SARS-CoV-2 transmission, modeling, and projected impacts.

